# Parental Anxiety in Differences of Sex Development and Hypospadias: A Psychosocial Characterization

**DOI:** 10.1101/2025.08.07.25332342

**Authors:** S Schlegel, J Kremen, A Dutcher, S Pereira, I Holm, YM Chan

**Affiliations:** Harvard Medical School; Boston Children’s Hospital; Boston Children’s Hospital; Baylor College of Medicine

## Abstract

**Introduction:** Parents of children with differences of sex development (DSD) and hypospadias experience significant psychological distress, but the specific factors contributing to anxiety in these parents remain poorly understood.

**Objectives:** Identify psychosocial factors associated with anxiety symptoms among parents of children with urogenital conditions and compare anxiety levels to parents in other pediatric care settings.

**Methods:** We conducted a cross-sectional survey study of 108 parents of children with urogenital conditions without established genetic diagnoses. Assessments included anxiety symptom scores (GAD-7), stigma measures, quality of life instruments, and demographic factors. Results were compared with 346 parents from BabySeq, a clinical trial of newborn genomic sequencing: 278 parents of healthy newborns, 68 parents of NICU infants. Backward elimination and LASSO regression modeling with multiple imputation identified key factors associated with anxiety symptoms.

**Results:** Clinically significant anxiety (GAD-7 ≥5) was present in 33% of parents of children with urogenital conditions, similar to the rate in parents of NICU infants (34%) and significantly higher than in parents of healthy newborns (21%, p=0.03). Forty-one percent of parents reported self-blame for their child’s condition, while 16% blamed their partners. DSD-related quality of life decreased with increasing stigma (p=0.001), genital atypicality (p=0.03), and anxiety scores (p=0.04). Four factors consistently predicted increased anxiety across multivariate models: higher intolerance of uncertainty, increased parental health-related stress, lower educational level, and greater emotional stigma. Additional factors included greater genital atypicality, social isolation, and self-blame.

**Discussion:** Parents of children with urogenital conditions experience anxiety levels comparable to those with critically ill infants despite their children being medically stable. The identification of specific risk factors, particularly uncertainty intolerance and emotional stigma, provides targets for clinical screening and early intervention to support these families.

## INTRODUCTION

When a child is diagnosed with a difference of sex development (DSD) or hypospadias, families face a unique set of medical, social, and psychological considerations that can significantly impact family dynamics and wellbeing. For these families, the experience often extends beyond medical management to encompass profound psychosocial dimensions that may persist throughout the child’s development.

Differences of sex development are generally defined as congenital conditions in which chromosomal, gonadal, or anatomical sex is atypical.^1,2^ This umbrella term encompasses a diverse spectrum of conditions, including sex-chromosome DSD, in which an individual has a sex chromosome complement other than XX or XY. Some clinicians include diagnoses such as Klinefelter syndrome and Turner syndrome within the DSD umbrella, although many researchers focus specifically on 46,XX patients with virilization and 46,XY patients with varying degrees of undervirilization.

Hypospadias, a condition in which the urethral meatus is positioned at a location other than the tip of the glans penis, represents one of the most common congenital anomalies of the male genitourinary system. While differences of sex development causing atypical genital development are relatively uncommon, occurring in approximately one out of every 4,500–5,500 births,^2^ hypospadias affects one out of every 150 to 300 male live births.^3,4^ This rate exceeds that of Down syndrome in the United States (one out of 667 live births),^5^ making hypospadias one of the most common congenital anomalies^5^ and underscoring the substantial public health significance of these urogenital conditions. The combined impact of these conditions creates a sizable population of families navigating a similar set of psychosocial challenges.

Research has consistently documented the profound psychological burden experienced by the parents of children with urogenital conditions such as hypospadias or DSD. One study found that the parents of children with DSD report posttraumatic stress levels similar to those of parents whose children have survived cancer.^6^ In addition, being the parent of a child with a urogenital condition such as hypospadias or a DSD can lead to significant anxiety, diminished quality of life, and feelings of stigmatization.^7,8^ When the cause of a child’s urogenital condition remains undiagnosed, this diagnostic uncertainty may worsen the psychological burden for parents.^9^ This psychosocial stress not only affects parental wellbeing, it also has the potential to create ripple effects throughout the family system. The possible intergenerational impact is particularly concerning, as children of anxious parents demonstrate a significantly higher risk of developing anxiety disorders themselves.^10–12^

Despite growing recognition of these psychosocial challenges, significant knowledge gaps persist in understanding the complex interplay of factors contributing to distress among caregivers of children with DSD. Recent guidance from a multi-stakeholder group emphasized the need to develop interventions promoting positive psychosocial adaptation and calling for more comprehensive research approaches in this area.^13^ In particular, international experts have raised concern over the use of single quality-of-life measures in prior studies of caregivers of children with DSD, which may not capture interactions between multiple psychosocial dimensions.^2^

This study addresses these knowledge gaps through a detailed assessment of psychosocial factors affecting parents of children with urogenital conditions, with particular attention to identifying specific contributors to clinically significant anxiety symptoms. By identifying factors that most strongly associate with parental distress, this research will help clinicians to screen for at-risk families and tailor interventions to support the psychological health of both parents and children facing these challenging diagnoses.

## METHODS

### Participants

We recruited the parents of patients with urogenital conditions without a known underlying genetic diagnosis from our hospital’s Urology, Endocrinology, and DSD specialty clinics as part of a broader study on the impact of genetic testing. In total, 108 parents completed baseline surveys. The sample size of 108 parents was based on recruitment feasibility within the study period. Patients who already had an established genetic diagnosis were excluded; thus, we excluded patients with sex chromosome mosaicism, virilizing congenital adrenal hyperplasia, and genetically confirmed diagnoses of androgen insensitivity syndrome. All parents were literate in English, Spanish, or Arabic (as validated study measures were not available in other languages).

### The BabySeq Project Dataset

To compare our baseline findings with another population in a similar region of the country and with similar sociodemographic characteristics, we analyzed data from the BabySeq Project. The BabySeq Project is a randomized clinical trial exploring the medical, behavioral, and economic impact of using whole-exome sequencing as an adjunct to traditional newborn screening in healthy newborns and infants in the neonatal intensive care unit (NICU).^14^ Infants were eligible to participate in the study if they were younger than 43 days, had not previously undergone exome sequencing, were born of a single-gestation pregnancy, and were either admitted to the Brigham and Women’s Hospital (BWH) Well Newborn Nursery or NICU or were admitted to the Boston Children’s Hospital (BCH) NICU or other ICU. Families were excluded if the parents were non-English speaking, younger than 18 years of age, or had impaired decisional capacity, or if clinical considerations precluded drawing 1 mL of blood from the infant.

### Survey Measures

The surveys analyzed in this study included questions that spanned multiple different psychosocial domains (Table 3). For the parents of children with urogenital conditions (“Urogenital cohort”), we conducted comprehensive psychosocial assessments using all domains listed in Table 3. For comparison with the BabySeq Project, we analyzed a subset of the measures collected in both studies: GAD-7 anxiety symptom scores, PHQ-9 depression symptom scores, and demographic variables (gender, race, ethnicity, age, and household income).

The “participant characteristics” category of questions included sociodemographics such as race, ethnicity, relationship status and household income, specific questions about the parent’s medical and family history, perceptions of genetic testing based on previous experiences (developed in collaboration with BabySeq investigators),^14,15^ and measures of attitudes and confidence such as the Intolerance of Uncertainty scale (to assess parents’ ability to cope with uncertainty).^16^ Multiple validated tools were included to measure the psychological wellness of parents and family units, such as the PHQ-9 scale to measure depressive symptoms,^17^ the GAD-7 scale to measure anxiety symptoms,^18^ the Parenting Stress Index to measure stresses in the parent-child relationship,^19,20^ the Child Health Worry Scale to measure the parent’s assessment of the child’s vulnerability,^21^ and the Multidimensional Scale of Perceived Social Support to measure the parent’s social connectedness.^22,23^ The surveys also included questions specific to patients undergoing genetic testing, such as questions developed in collaboration with BabySeq investigators about self-blame and partner blame regarding the child’s diagnosis. Questions specific to patients with urogenital conditions included the DSD-Associated Stigma measure, a validated instrument used to assess stigma in DSD patients.^24^ Finally, we included the DSD Health-Related Quality of Life instrument, a validated tool that includes questions about topics such as medical decision-making, the child’s role in family activities, gender concerns, social functioning, concerns about the future, and experiences with the healthcare system.^25,26 27,28^The surveys were administered online using REDCap,^26^ with paper versions offered to those unable to complete electronic forms.

### Multiple Imputation

To address missing survey data (6.2% of summary-score data), we performed multiple imputation using the Multiple Imputation by Chained Equations (MICE) package in R. This package acknowledges the uncertainty of the missing values by creating multiple plausible estimates for the missing values rather than using listwise deletion, which has the potential to introduce bias.^27^ Following recommended guidelines that the number of imputations should be greater than the percentage of missing data,^27,28^ we conservatively performed 20 iterations of imputation.

### Measurement of Genital Typicality

To quantify the patients’ degree of genital atypicality, we reviewed medical records and used findings from physical examination to determine the External Genitalia Score (EGS), a standardized description of external genitalia for preterm and term babies up to 24 months of age.^29^ The EGS gives points for different physical features, such as length of the genital tubercle, location of the urethral meatus, degree of fusion of the labioscrotal structures, and ability to palpate the gonads. Possible scores range from typical female appearance (0) to typical male appearance (12), with an EGS of 6 describing the most atypical appearance.^29^

In this study, if a range of possible scores was suggested by the exam documented in the chart, the final score selected was the average of the highest and lowest possible scores consistent with the exam. Based on the hypothesis that psychosocial stressors would be associated with degree of atypicality rather than whether the child’s genitalia appeared more typically male or typically female, we also calculated a “genital typicality score” by taking the absolute value of the difference between the child’s EGS and 6 (the most atypical, intermediate score).

### Statistical Analyses

#### Mixed-Effects Bivariate Modeling

To understand the psychosocial factors that contribute to the quality of life of the families of children with urogenital conditions, we performed association studies between DSD-Related Quality of Life (QOL) and other psychosocial tools in the survey. To account for the possibility of a correlation between the responses for the mother and father of the same child, we created mixed-effects models for this analysis. This technique accounts for potential correlations between parent responses by incorporating a random-effect value for each parent pair.

We also used linear and logistic mixed-effects models with the GAD-7 score as the outcome to test our hypothesis that the degree of anxious symptoms (as measured by GAD-7 scores) in the parents of children with urogenital conditions was greater than that experienced by the parents of children in the BabySeq healthy newborn group but similar to that experienced by the parents of children in the BabySeq NICU group. We added additional psychosocial variables to the mixed-effects models to control for possible confounders of group differences in anxiety scores, such as the parent’s gender, ethnicity, race, age, and household income. Clinically significant anxiety symptoms were defined as GAD-7 scores ≥5, consistent with established clinical cutoffs.^30^

#### Multivariate Modeling of GAD-7

To elucidate the major psychosocial features related to the high levels of anxious symptoms in the parents of children with urogenital conditions, we performed multivariate modeling of GAD-7 scores. To determine which variables should be included in the multivariate model, we first examined relationships between variables using correlation tables, using Pearson correlation coefficients for variables with minimal correlation between parent pairs and using the square root of the R^2^ value of linear mixed-effects models for variables demonstrating inter-parental correlation. Variables with correlation coefficients >0.7 or <-0.7 were considered for exclusion to prevent collinearity. Only one such pair was identified (r=0.87); the variables had nearly identical values for all participants, as both described a similar phenotype (“presence of hypospadias with urethral opening within the scrotum”), and we retained only the variable with the stronger association with GAD-7.

For measures with subdomains, we excluded aggregate psychosocial scores in favor of including their subscores as more specific predictors. We also excluded variables where most participants gave the same response (i.e., variables that did not meaningfully differentiate between participants). Finally, because PHQ-9 (for depressive symptoms) is known to be closely associated with GAD-7 and we were concerned that its inclusion would mask the contributions of other variables to the GAD-7 score, we also excluded PHQ-9 and other depression scores in our main analysis models.

To build the multivariate models, we employed the Least Absolute Shrinkage and Selection Operator (LASSO) technique, which reduces the risk of overfitting through cross-validation and coefficient shrinkage. This approach is superior to many other methods used for multivariate regression modeling due to this reduced risk of overfitting, increased model interpretability, and improved ability to accurately model the same outcome in other samples drawn from the same population.^31^

To address missing data, we applied the LASSO technique to our multiply imputed dataset using the “separate approach” described by a recent publication,^28^ which constructs a different model for the dataset arising from each imputation iteration. We selected this method due to its more parsimonious selection process and increased predictive performance. Because there was minimal correlation between GAD-7 scores within parent pairs (r = 0.08), we did not include random effects for parent clustering, similar to the approach used in the BabySeq Project when there was no significant within-family clustering in the outcome measure.^15^

We implemented 10-fold cross-validation, with lambda values derived by averaging the optimal values across all 20 imputations. The variable coefficient outputs for each imputed dataset were then averaged (if a particular variable was not selected in one of the imputations, then that variable was considered to have a coefficient of zero).

We constructed two models: LASSO Model 1 included all variables except those explicitly excluded above. Following established practice for data analysis when the number of predictors exceeds sample size, we also constructed LASSO Model 2, for which we first identified variables that were associated with GAD-7 in univariate analysis with *p* ≤0.15 before applying LASSO.^32^

To compare the relative contributions of different variables, normalized predictor variable coefficients were calculated by taking the absolute value of the mean of all normalized predictor coefficients from iterations where each predictor was included in the model. Finally, we cross-checked our models by repeating the model-building on the multiply imputed dataset with the backward selection technique (a method that removes variables one by one based on their pooled *p*-values in the presence of all other variables).^33^

## RESULTS

### Participant Demographics

Our study included 108 parents of children with urogenital conditions, with 55% mothers and 45% fathers (median age 36 years, range 25-56). Most participants (80%) described their race as white, with a smaller proportion describing themselves as Asian (11%), Black or African American (6%), or more than one race (3%). Twenty-nine percent of parents had a household income of at least $200,000/year, and 13% reported an income of less than $50,000/year. The age of the patients in this study ranged from 0-15 years, with a median age of 2 years. The working diagnoses of the patients at the time of study enrollment included isolated hypospadias (59%), hypospadias with undescended testicles and/or micropenis (21%), conditions affecting testicular development (6%), conditions affecting androgen synthesis/action (5%), testicular regression syndrome (6%), and androgen excess (2%). Complete demographics for patients and parents are presented in Tables 1 and 2, respectively.

**Table 1.** Patient Demographics.

| <u>Working Diagnosis at Time of Genetic Testing</u> | ***n*** | **%** |
| --- | --- | --- |
| XY |  |  |
| Condition Affecting Testicular Development | 7 | 6 |
| Condition Affecting Androgen Synthesis/Action | 5 | 5 |
| Hypospadias with Undescended Testicles/Micropenis | 23 | 21 |
| Isolated Hypospadias | 64 | 59 |
| Testicular Regression Syndrome | 7 | 6 |
| XX |  |  |
| Androgen Excess | 2 | 2 |
| <u>External Genitalia Score</u> | **Median** | **Range** |
| Children Reared as Girls (n=4) | 2 | 0-5.5 |
| Children Reared as Boys (n=98) | 10.1 | 4.75-12 |
| <u>Age (years)</u> | 2 | 0-15 |

**Table 2.** Parent Demographics in Urogenital and BabySeq Cohorts.

|  | **Urogenital** | **Well Baby** | **NICU Baby** |
| --- | --- | --- | --- |
| Parent Gender | **n(%)** | **n(%)** | **n(%)** |
| Female | 57(55) | 159(57) | 34(50) |
| Male | 46(45) | 119(43) | 34(50) |
| Ethnicity |  |  |  |
| Hispanic | 10(10) | 16(7) | 0(0) |
| Non-Hispanic | 87(90) | 221(93) | 66(100) |
| Race |  |  |  |
| Asian | 11(11) | 35(14) | 5(8) |
| Black/African American | 6(6) | 5(2) | 0(0) |
| White/Caucasian | 78(80) | 206(81) | 58(88) |
| More than One Race or Other | 3(3) | 8(3) | 3(5) |
| Relationship Status |  |  |  |
| Single and Never Married | 3(3) | 2(1) | 0(0) |
| Single and Separated or Divorced | 1(1) | 0(0) | 0(0) |
| In a Committed Relationship | 101(96) | 247(99) | 63(100) |
| Partner is Child's Biological Parent | 92(93) | 240(98) | 61(97) |
| Household Income |  |  |  |
| <\$20,000 | 5(5) | 2(1) | 1(2) |
| \$20,000-\$49,999 | 8(8) | 3(1) | 1(2) |
| \$50,000-\$99,999 | 15(15) | 33(13) | 12(20) |
| \$100,000-\$199,999 | 42(42) | 101(41) | 28(46) |
| \$200,000-\$499,999 | 27(27) | 95(39) | 16(26) |
| \$500,000 or above | 2(2) | 11(4) | 3(5) |
| Highest Completed Education |  |  |  |
| 8<sup>th</sup> Grade or less | 1(1) | 0(0) | 0(0) |
| 9<sup>th</sup> to 12<sup>th</sup> Grade or GED | 7(7) | 2(1) | 1(2) |
| Vocational School or Some College | 13(12) | 15(6) | 7(11) |
| College Graduate | 40(37) | 81(33) | 21(33) |
| Graduate or Professional School | 46(43) | 151(61) | 34(54) |
| Partner's Highest Completed Education |  |  |  |
| 8<sup>th</sup> Grade or Less | 4(4) | NM | NM |
| 9<sup>th</sup> to 12<sup>th</sup> Grade | 9(9) | NM | NM |
| Vocational School or Some College | 14(13) | NM | NM |
| College Graduate | 33(31) | NM | NM |
| Graduate or Professional School | 45(43) | NM | NM |
|  | **Mean (Range)** |  |  |
| Age (years) | 36(25-56) | 34.5(22-53) | 33(23-49) |

Not all participants answered all questions, so percentages and totals reflect those who answered the relevant questions.
NM = "Not Measured" in survey.

**Table 3.** Psychosocial Domains Measured in Baseline Surveys.

| Variable Domains | Survey Tools |
| --- | --- |
| **Participant Characteristics** |  |
| Socio-demographics | Sociodemographic questionnaire derived from the NHANES Demographic & Background questionnaire, as well as NIH standards for reporting race/ethnicity |
| Genetic Perception | Survey instruments developed in collaboration with BabySeq investigators to describe previous experiences with genetic testing<sup>14,15</sup> |
| Supports | Instrument derived from Pew Research Center to assess religion and spirituality;<sup>66</sup> The Multidimensional Scale of Perceived Social Support<sup>22,23</sup> |
| Confidence | The Intolerance of Uncertainty scale<sup>16</sup> |
| Personal and Family History | Tools used to gather health history derived from the short Form Health Survey (SF-12),<sup>67</sup> the Pregnancy Risk Assessment Monitoring System,<sup>68</sup> and the BabySeq Project;<sup>14,15</sup> questions about fertility and DSD diagnosis developed by the Boston Children's Division of Endocrinology's Disorders of Sex Development Research Conference |
| **Psychological Wellness** |  |
| Personal Distress (Depression & Anxiety) | The PHQ-9 scale<sup>17</sup> and the GAD-7 scale<sup>18</sup> |
| Parent-Child Relationship and Child-Centered Stress | The Parenting Stress Index version 4<sup>19,20</sup> |
| Perceptions of Child | The Child Health Worry Scale derived from the Vulnerable Baby Scale<sup>21</sup> |
| Perceived Self-Blame, Partner-Blame | Instrument developed in collaboration with BabySeq investigators as a measure of blame regarding child's diagnosis<sup>14,15</sup> |
| **DSD-Specific Measures** |  |
| DSD-Associated Stigma | The DSD-associated Stigma measure<sup>24</sup> |
| DSD-Associated Quality of Life | The DSD Health-Related Quality of Life instrument<sup>25</sup> |

When comparing the Urogenital cohort with the BabySeq cohorts, we found that a greater percentage of the Urogenital cohort had household income <$50,000 per year compared with the parents of healthy infants in the newborn nursery (“Well Baby Cohort,” N = 278) and the parents of ill newborns in the NICU (“NICU Baby Cohort,” N = 68) (13%; 2%; 3%), and a lower percentage had graduated from college compared with the BabySeq Well and NICU cohorts (80%; 93%; 87%).

### Parental Self-Blame and Partner Blame

Because previous studies identified self-blame as a significant cause of stress in the parents of children with genetic conditions,^34^ we examined participants’ experiences of blame. We found that 41% of participants described blaming themselves at least “a little” for passing harmful genes to their children (Figure 1), similar to the 42% frequency of self-blame that has been reported by parents of children with cystic fibrosis.^34^ In addition, 16% described blaming their partners at least “a little” (Figure 1).

**Figure 1.**
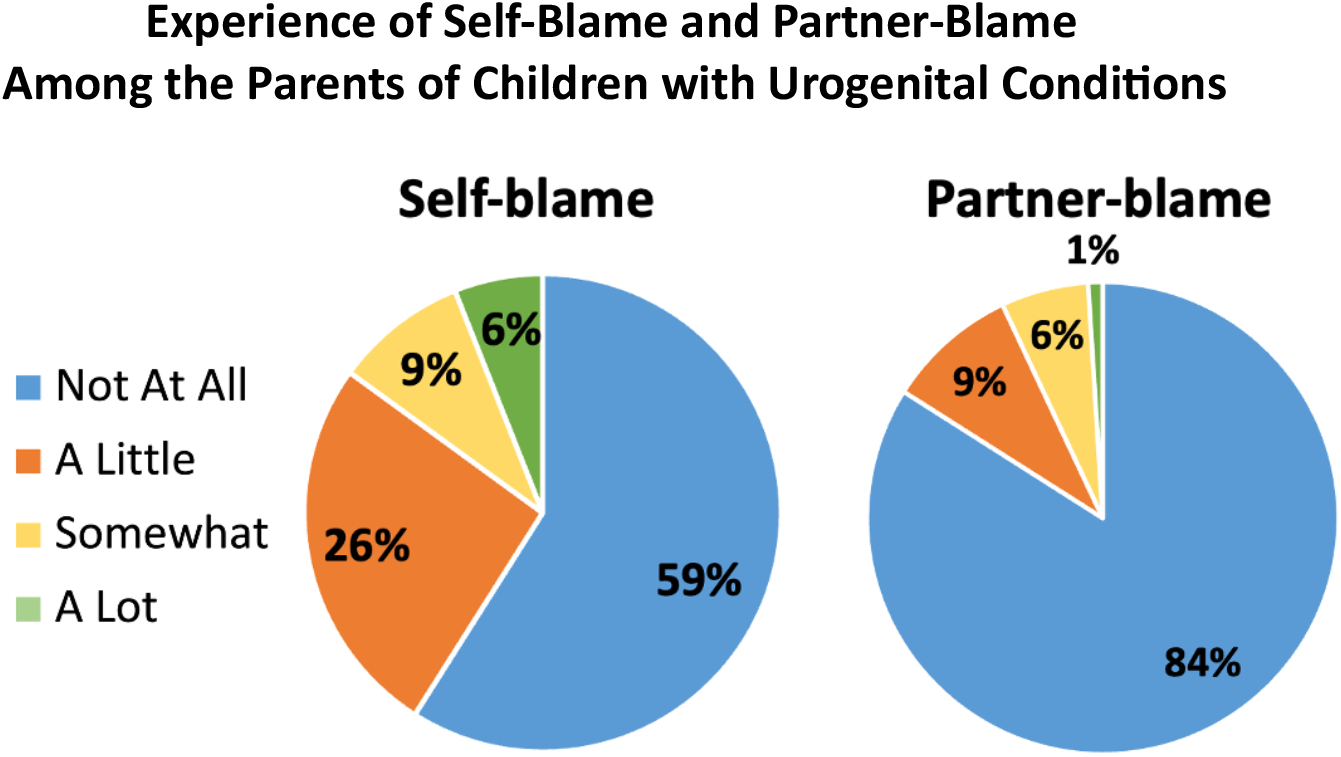
Responses to the survey question, “How much do you blame yourself/your partner for passing potentially harmful genes on to your child?”

### DSD-Related Quality of Life

To understand the psychosocial factors that contribute to diminished quality of life experienced by some families of children with urogenital conditions, we performed association studies between DSD-Related Quality of Life and other psychosocial tools in the survey using linear mixed-effects models. With this modeling technique, we found that DSD-related Quality of Life decreased with increasing DSD stigma (p=0.001), increasing genital atypicality (^29^p = 0.03; Figure 2), and increasing anxiety scores (p = 0.04; Figure 3). To further explore the presence of anxious symptoms in our population, given their association with DSD-Related Quality of Life and their previously identified association with having a child with a urogenital condition,^7^ we quantified the prevalence of anxious symptoms in our cohort. We found that 33% of respondents had GAD-7 scores demonstrating anxious symptoms in the clinically significant range, a greater proportion than the 16% frequency reported in the general population.^35^

**Figure 2.**
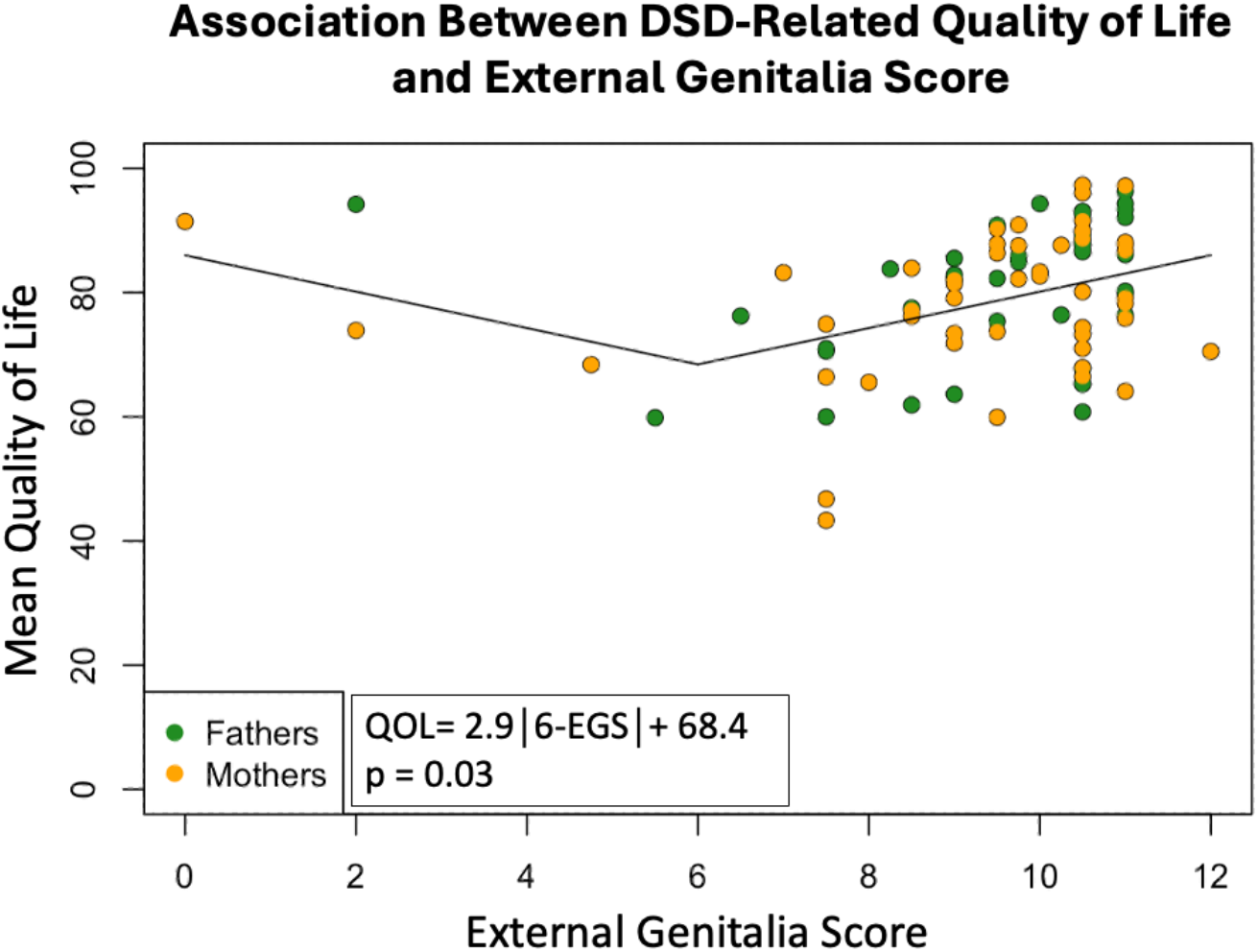
DSD-related Quality of Life decreases with increasing genital atypicality as measured by the External Genitalia Score (EGS). The EGS ranges from most typically female (0) to most typically male (12), with an EGS of 6 describing the most atypical appearance. For this analysis, linear mixed-effects modeling was performed with multiply imputed data. The data points in the graph represent only the data collected in the study and do not include the imputed values.

**Figure 3.**
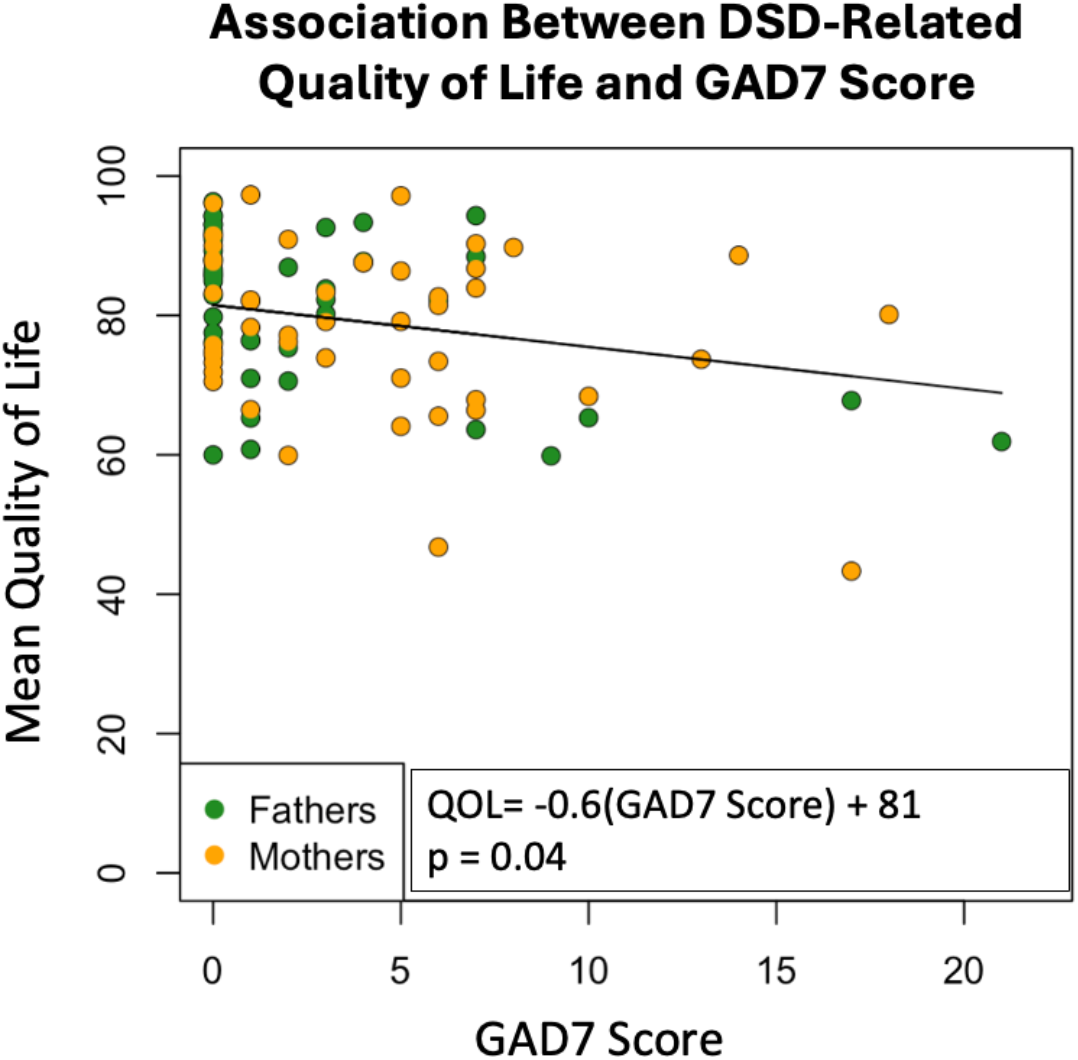
DSD-related Quality of Life decreases with increasing anxious symptoms as measured by the GAD-7 score (p = 0.04). For this analysis, linear mixed-effects modeling was performed with multiply imputed data. The data points in the graph represent only the data collected in the study and do not include the imputed values.

### Comparisons with the BabySeq Project

The Urogenital cohort had higher GAD-7 scores than parents of healthy newborns (p=0.007) (data not shown), even after controlling for parents’ gender, race, ethnicity, age, and household income (p=0.01), but similar GAD-7 scores to parents of children in the NICU (p=0.9).

Anxiety symptom scores in the clinically significant range were present in a greater proportion of the parents of children with urogenital conditions (33%) than the BabySeq healthy newborn cohort (21%; p = 0.03), but at a similar proportion to the BabySeq NICU cohort (34%; p=0.8), despite the fact that the children with urogenital conditions were clinically stable at home rather than in a critical care unit (Figure 4). After controlling for parents’ gender, race, ethnicity, age, and household income, the difference in the frequency of clinically significant anxiety between the DSD and Well groups remained borderline significant (p=0.051).

**Figure 4.**
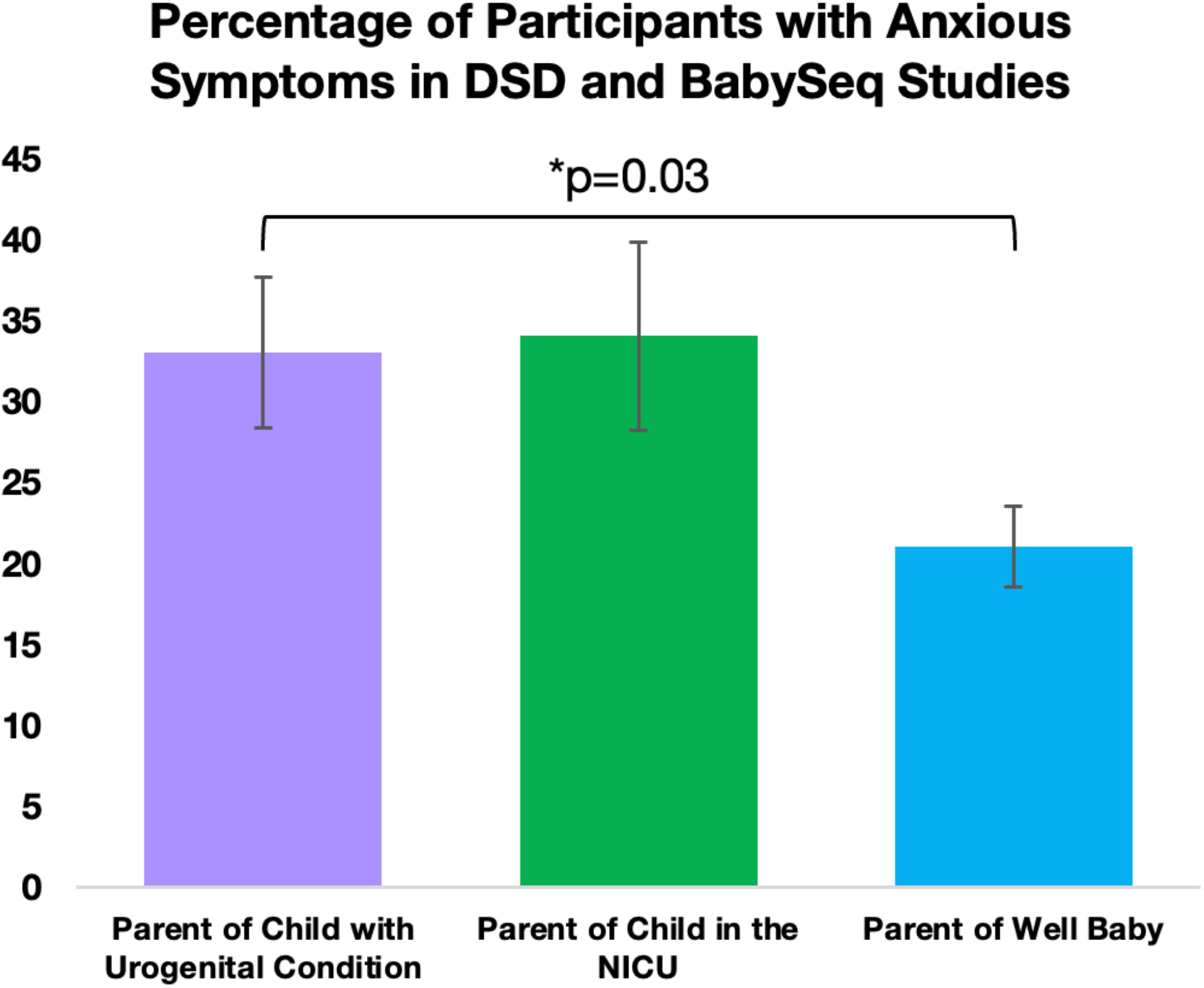
Anxious symptoms in the clinically significant range (GAD-7 score ≥5) were present in a greater proportion of the parents of children with urogenital conditions than in the parents of children in the BabySeq healthy newborn group. A comparable proportion of the parents of children in the BabySeq NICU group had clinically significant anxious symptoms. Standard error bars are shown. Linear mixed-effects modeling was used for the analysis.

There was no significant difference in the PHQ-9 scores for depressive symptoms of parents or the frequencies of clinically significant depression in the DSD, Healthy, and NICU cohorts, even when controlling for demographic factors (data not shown).

### Multivariate modeling of GAD-7 Scores

To elucidate the major psychosocial features related to the high levels of anxiety symptoms in the parents of children with urogenital conditions, we used a systematic modeling approach. Based on findings in other populations, we hypothesized that the high levels of anxiety symptoms observed among the parents of children with urogenital conditions would be most closely associated with other psychosocial issues such as increased depressive symptoms,^36^ other life factors such as poorer caregiver health,^37,38^ and socioeconomic factors such as lower income.^37^

To identify factors with the most robust associations with parental anxiety, we employed multiple complementary analytical approaches: backward elimination (which systematically removes less important variables) and LASSO regression applied to multiply imputed datasets (which created 20 separate models to account for missing data uncertainty, ensuring that our findings are not dependent on assumptions about missing values). Across all modeling approaches, four factors were consistently associated with increased anxiety: (1) higher intolerance of uncertainty, (2) increased parental health-related stress, (3) lower parental educational level, and (4) greater parental emotional stigma related to the condition (Figure 5). These four key factors all had mean coefficients that were among the 11 largest in LASSO Model 1 (built with all variables) and among the 8 largest in LASSO Model 2 (built with variables that were associated with GAD-7 in univariate analysis).

**Figure 5.**
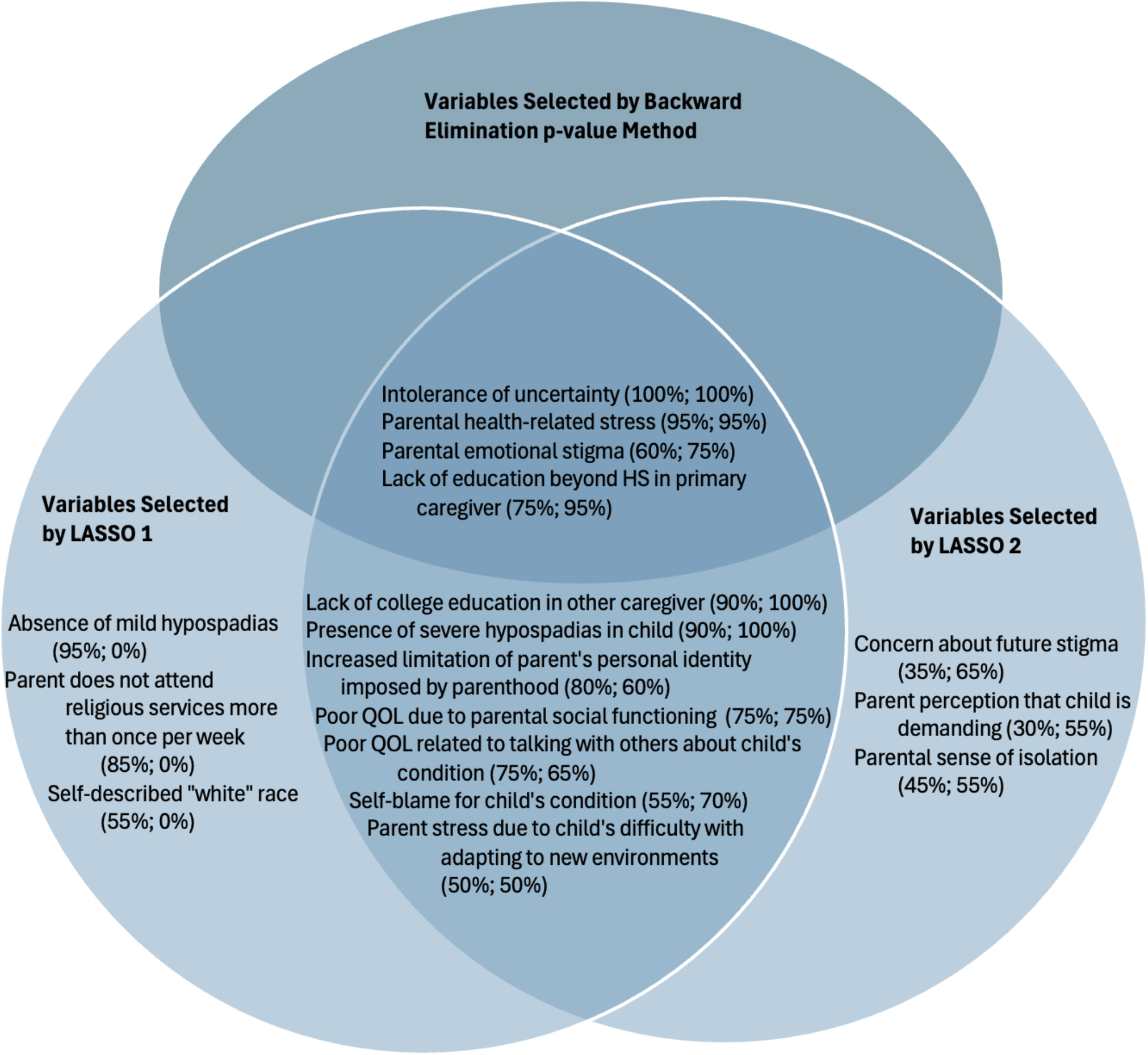
Variables Selected by Different Methods of Multivariate Modeling. LASSO 1 included as inputs all survey variables from the parents of children with urogenital conditions; LASSO 2 included only variables associated with GAD-7 with p ≤ 0.15. Variables shown in LASSO results were selected in at least 50% of all imputations from the multiply imputed datasets. Percentages describe the proportion of the 20 total LASSO analyses (one for each imputation) that identified the given variable as being associated with the parent’s GAD-7 score; the first percentage describes the results of LASSO 1, and the second percentage describes the results of LASSO 2. All variables selected by the backward elimination pooled p-value method were selected by both LASSO models. QOL: DSD-Associated Quality of Life. LASSO: Least Absolute Shrinkage and Selection Operator. HS: High School.

Other factors that frequently emerged in LASSO Model 1 and LASSO Model 2 included increased genital atypicality, loss of personal identity beyond parenting, poor parental social functioning, increased difficulty discussing the child’s condition with others, greater self-blame, and increased concerns about future stigma for the child, while frequent religious service attendance emerged as potentially protective.

Because PHQ-9 scores are known to correlate strongly with GAD-7 scores, we repeated our analyses including PHQ-9 to test the robustness of our findings. All four key factors remained significantly correlated with anxiety in these models, confirming their independent importance (Figure S1).

## DISCUSSION

In this study, we characterized experiences of stigma, quality of life, and self-blame among parents of children with congenital urogenital conditions (DSD and hypospadias) and identified key psychosocial factors associated with parents’ anxiety symptoms. Our findings highlight several important patterns that carry direct clinical implications for supporting these families.

First, we found high rates of parental self-blame. The prevalence of self-blame is similar to reports of self-blame among parents of children with cystic fibrosis^34^ and provides important insight into a potential driver of psychological distress in these families.

Our quality-of-life analyses revealed that increased experiences of stigma and greater genital atypicality were both associated with decreased quality of life among parents. While causality cannot be established in this cross-sectional study, these relationships highlight the impact of both social and physical factors in parental adjustment to these conditions.

Strikingly, we found that parental anxiety in this population is comparable to that experienced by parents of critically ill infants in the NICU. The similarity in anxiety levels suggests that having a child with a urogenital condition can be as psychologically distressing for parents as having a child in an intensive care unit, indicating that the emotional impact of these diagnoses should not be underestimated, and suggesting that psychological support should be as readily available in outpatient urogenital care as it is in acute critical-care settings.

Through multivariate modeling, we identified four key factors associated with increased anxiety in this population: increased parental health-related stress, lower parental educational level, greater emotional stigma related to the condition, and higher intolerance of uncertainty. Some of these factors – particularly poor parental health and lower educational level – likely contribute to anxiety in many populations beyond parents of children with urogenital conditions. However, the emotional stigma component appears uniquely relevant to this specific population, underscoring the distinct psychological burden these parents face. Our finding that emotional stigma strongly correlates with anxiety confirms the well-documented psychosocial impact of perceived societal attitudes toward urogenital conditions, particularly amidst increased public discourse around gender and biological sex. This finding adds to the established evidence that among the parents of children with DSD, stigma significantly contributes to the psychological burden for families,^39–41^ similar to patterns observed in other stigmatized conditions such as HIV and epilepsy.^42,43^

Uncertainty intolerance showed a remarkably strong association with anxiety in our population, remaining significant across all our analytical approaches – even when we controlled for depression symptoms (PHQ-9). The prominence of uncertainty intolerance in our model suggests that parents with higher anxiety may find it particularly challenging to cope with the unknowns regarding their child’s diagnostic workup, treatment outcomes, and future psychosocial adjustment to the urogenital condition. This finding is consistent with prior reports that illness uncertainty among the parents of children with DSD is comparable to that of parents of children with other chronic illnesses such as type 1 diabetes mellitus,^7^ and previous research has found that illness uncertainty in such chronic conditions is a robust predictor of parental psychological distress over time.^44^

Social identity concerns also emerged as a prominent theme in our models. Notably, the PSI4 subscale “increased limitation of parent’s personal identity imposed by parenthood” appeared frequently in both LASSO models. This experience – where parents feel they must devote most of their energy and identity to parenting at the expense of their individual identity – has been well-documented among parents of children with various chronic conditions.^43,45^ The prominence of the PSI4 subscale “parent perception that child is demanding” in our models further reflects the heightened caregiving burden associated with managing a child’s chronic condition.

Beyond individual parenting experiences, broader concerns about social functioning also emerged in our analyses. The “parental sense of isolation” subscale consistently emerged alongside other social factors such as “poor QOL due to parental social functioning” and difficulties discussing the child’s condition with others.

These findings align with research documenting social isolation among parents of children with other chronic conditions, including disabilities,^46^ inflammatory bowel disease,^47^ ADHD,^48^ and autism spectrum disorder.^49^ The potentially protective effect of religious service attendance against anxiety identified in our models further suggests the importance of community integration.

### Clinical Recommendations

Our findings suggest several important clinical practice changes to better support parents of children with urogenital conditions. First, we recommend systematic screening for potentially modifiable factors associated with anxiety, particularly intolerance of uncertainty and emotional stigma (see intervention approaches discussed below). This targeted screening would help identify parents who may benefit from early psychosocial intervention. Screening for parental health concerns is also important, as our findings emphasize that a child’s urogenital condition represents only one aspect of a family’s overall experience, and other stressors may compound psychological impact. Parents who screen positively for these risk factors should undergo formal anxiety assessment and may benefit from proactive referral to appropriate mental health services.

Given the high prevalence of self-blame in this population, clinicians should address this topic during clinical visits. This discussion should normalize these feelings while providing accurate information about inheritance patterns. Clinicians should also provide guidance on approaches to navigating social situations and sharing information about the child’s condition, addressing the social isolation many parents experience.

Our finding that 33% of parents experience clinically significant anxiety – a rate comparable to parents with children in intensive care units – provides strong justification for embedding mental health services within urogenital specialty care settings. These services could include specialized support groups focused on strategies for coping with uncertainty and managing stigma. Research in other populations offers promising approaches: studies among oncology patients and caregivers have demonstrated success with home-based interventions that provide support around illness uncertainty by teaching skills to obtain information, increase one’s assertiveness, and learn to live with uncertainty.^50–52^ To address stigma, evidence in conditions such as HIV/AIDS, mental illness, and epilepsy, supports multiple approaches including individual counseling/cognitive behavioral therapy, advocacy and support groups, community-based programs, and broader community education initiatives.^53,54^

Several existing interventions to improve parental anxiety also show promise for adaptation to this population. For example, an online mindful parenting intervention demonstrated medium effect sizes for improving symptoms of depression and anxiety, with additional benefits for parenting discipline, self-compassion, and child emotional reactivity. This intervention also showed specific improvements in parental role restriction – a factor prominently identified in our models.^55^

Finally, the protective effect of religious service attendance suggests the value of community integration. Clinicians should inquire about and encourage connection to supportive community resources as potential buffers against isolation and anxiety. This aligns with our finding that social factors play a crucial role in parental well-being, highlighting the importance of addressing not only medical aspects of care but also the broader social context in which families navigate these conditions.

### Study Strengths

Our study has several notable strengths. First, the extensive nature of our psychosocial assessment, incorporating multiple validated instruments across various domains, allowed us to capture a detailed picture of parental experiences. The inclusion of both mothers and fathers provides valuable insight into the experiences of both parents, addressing a gap in previous literature that often focused primarily on maternal perspectives.

The comparison with BabySeq cohorts represents another key strength, allowing us to contextualize our findings within both healthy and acute-care populations. This comparison provides important insights into the relative psychological impact of having a child with a urogenital condition compared with other medical scenarios.

Our analytical approach also represents a methodological strength. The use of multiple imputation and state-of-the-art modeling techniques, including LASSO variable selection, enhanced the robustness of our findings. Additionally, the consistency of our findings across different analytical approaches supports their validity.

Lastly, our study’s focus on parents of children without established genetic diagnoses addresses an important knowledge gap, as this population may face unique uncertainties and challenges compared to families with confirmed genetic conditions.

### Study Limitations

Several limitations should be considered when interpreting our results. The diversity of diagnoses in our study population limited our ability to identify diagnosis-specific patterns, allowing only for analysis of patterns applicable across multiple urogenital conditions. Our sample also demonstrated limited demographic diversity, with most participants identifying as white, high-income, and highly educated. The study design was also subject to potential recall bias in retrospective questions and responder bias. Missing data is a limitation of any survey-based methodology, although we attempted to mitigate the effects of missing data through use of multiple imputation.

### Future Directions

Several important research directions emerge from this work. One notable stressor for families of children with urogenital conditions is the discomforting uncertainty regarding the underlying cause of the condition. Genetic testing plays an important role in the evaluation and care of individuals with urogenital conditions and has the potential to decrease these feelings of uncertainty. However, research on the impact of receiving genetic testing results is mixed: some studies found that receiving genetic testing results may be associated with increased anxiety in parents,^56–58^ but others observed a decrease in anxiety scores after receiving positive genetic testing results^15,59^ and a feeling of relief from worry after receiving exome-sequencing results.^60^ To provide sensitive care for children with urogenital conditions, it is crucial to understand the psychological consequences of pursuing genetic testing.

Future comparative analysis of anxiety correlates between our cohort and the BabySeq cohorts may also help to distinguish which associations are specific to parents of children with urogenital conditions versus common to all anxious parents.

Finally, we suggest that future studies explore how anxiety and support needs evolve as children age and examine the impact of protective factors like community involvement through qualitative research. Given our findings regarding social isolation and stigma, interventions targeting these domains warrant investigation. Particularly promising are narrative medicine and reflective writing interventions, which have shown benefits for psychosocial stress in caregivers^61–63^ and in individuals experiencing high levels of shame or stigma.^64,65^

## Summary and Conclusions

Our findings contribute valuable insights into the psychological experiences of parents of children with urogenital conditions. The discovery that these parents experience anxiety levels comparable to those with children in intensive care settings underscores the profound psychological impact of these conditions. The four key anxiety-related factors we identified – uncertainty intolerance, parental health challenges, educational factors, and emotional stigma – provide a framework for understanding parental distress in this population. Particularly notable is how these factors interact with social experiences, illuminating the isolation many families experience while managing both medical complexities and social challenges. The potential protective effect of community engagement through religious attendance offers an intriguing direction for further exploration of resilience factors.

This work contributes to the growing body of urogenital care research, moving beyond purely medical considerations toward a more comprehensive understanding of family experiences. By documenting both challenges and potentially protective factors, we establish a foundation for future improvements in supportive care that addresses the full spectrum of needs these families face. Our findings suggest that an integrated approach to care – one that acknowledges both physical and psychosocial dimensions – may ultimately enhance both parental wellbeing and child outcomes in this population.

## Data Availability

All data produced in the present study are available upon reasonable request to the authors.

## Acknowledgments

Carter Petty, Malcolm Matheson, Saakshi Daswani

## Trial Registration

ClinicalTrials.gov identifier NCT03102554

