## Supplemental Figure S1 for "Parental Anxiety in Differences of Sex Development and Hypospadias: A Psychosocial Characterization"

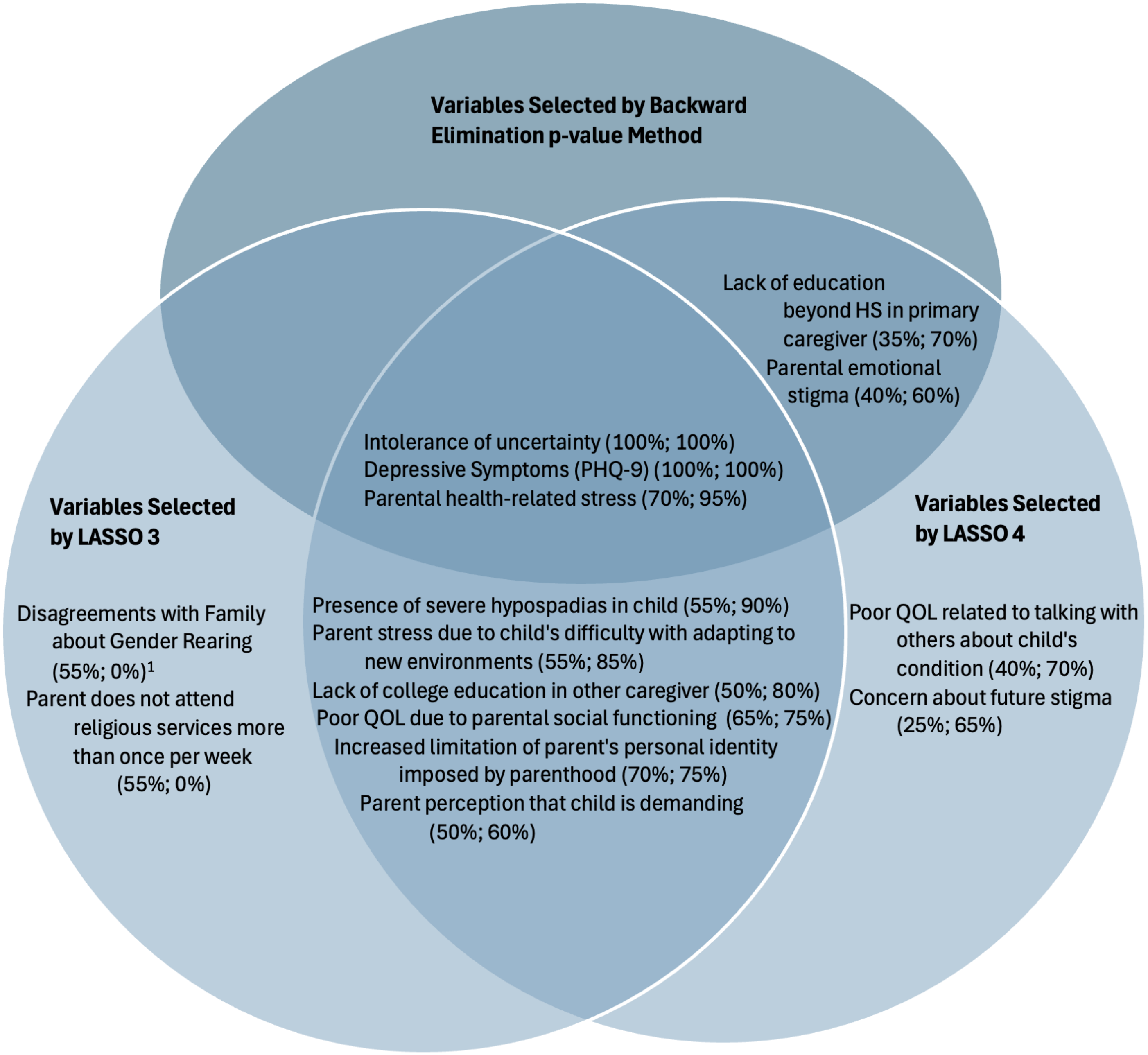


**Figure S1. Variables Selected by Different Methods of Multivariate Modeling in the Presence of the PHQ-9 Depression Scale.** LASSO 3 included as inputs all survey variables from the parents of children with urogenital conditions including PHQ-9; LASSO 4 included only variables associated with GAD-7 with $p \leq 0.15$, including PHQ-9. Variables shown in LASSO results were selected in at least 50% of all imputations from the multiply imputed datasets. Percentages describe the proportion of the 20 total LASSO analyses (one for each imputation) that identified the given variable as being associated with the parent's GAD-7 score; the first percentage describes the results of LASSO 3, and the second percentage describes the results of LASSO 4. All variables selected by the backward elimination p-value method were selected by LASSO 4. All 4 key variables from Figure 5 were still selected in at least one LASSO model, and 2 key variables (higher intolerance of uncertainty and increased parental health-related stress) were selected in both LASSO models. The following variables had been selected by LASSO 1 or LASSO 2, but were not selected in the presence of PHQ-9: Self-blame for child's condition, Parental sense of isolation, Self-described "white" race, Absence of mild hypospadias. [1]Variable that was not selected in LASSO 1 or LASSO 2. QOL: DSD-Associated Quality of Life. LASSO: Least Absolute Shrinkage and Selection Operator. HS: High School.
